# Beyond Awareness: Factors Associated with Willingness to Participate in Living Kidney Donation

**DOI:** 10.64898/2026.09.22.26363701

**Authors:** Shreya Madan, Elizabeth M Thomas

## Abstract

**Background:** Living donor kidney transplantation improves outcomes for eligible patients with kidney failure, yet living donation remains limited. Public awareness about the possibility of living donation may not be sufficient to translate into willingness to donate. This study evaluated knowledge, attitudes, personal exposure, perceived barriers, motivators, and financial considerations associated with willingness to participate in living kidney donation among adults in San Antonio, Texas.

**Methods:** A cross-sectional community-based survey was conducted among 346 adults. The survey assessed demographic characteristics, knowledge about kidney transplantation and living donation, personal exposure to dialysis, transplantation, and living kidney donation, misconceptions regarding donation, motivating and discouraging factors, financial considerations, and willingness to become a living donor. Associations with willingness were evaluated using multivariable logistic regression models.

**Results:** Although 83.8% of participants knew that a healthy person could donate one kidney while living, only 68 participants (19.8%) expressed general willingness to become a living donor. Only 40.0% identified kidney transplantation as the best treatment for eligible patients with kidney failure. Demographic and socioeconomic factors were not independently associated with willingness, and individual misconceptions were not independently associated with willingness. In the integrated model, knowing a living kidney donor was associated with greater willingness (OR 2.82, 95% CI 1.33–5.95), whereas knowing a transplant recipient was associated with lower willingness (OR 0.42, 95% CI 0.19– 0.96). Helping someone in need was associated with greater willingness (OR 2.48, 95% CI 1.39–4.44), while concern about long-term donor health complications was associated with lesser willingness to donate (OR 0.48, 95% CI 0.26–0.90). Nearly half of respondents who answered the compensation question selected a monetary amount that could motivate donation.

**Conclusions:** In this community-based sample, awareness that living donation is possible did not translate into broad willingness to donate. Willingness appeared to be shaped by personal exposure, altruistic motivation, donor safety concerns and financial considerations. Community education should move beyond basic awareness and address donor safety, financial protection, living donor experiences, and the benefits of kidney transplantation.

## Introduction

Chronic kidney disease (CKD) affects an estimated 35.5 million adults in the United States [1], with a growing number progressing to end-stage renal disease (ESRD) requiring dialysis or kidney transplantation. Kidney transplantation, particularly from living donors, is associated with better patient and graft survival compared to dialysis or deceased donor transplantation [2,3]. Despite these benefits, living donor transplantation accounts for less than a quarter of kidney transplants performed annually in the United States [3], suggesting that barriers to donation exist long before potential donors enter formal evaluation pathways.

Several studies have collectively improved our understanding of living kidney donation from the perspectives of the public [4,5], donor candidates [6,7], and actual donors [8]. Still, most of these studies have focused on specific domains such as financial barriers [4,6], donor safety concerns [4], and socioeconomic status, geography, perceptions of fairness in the transplant system, and basic awareness of living donation [5]. Additionally, some national surveys have evaluated willingness to donate primarily in the context of donation to a family member, a circumstance in which an individual’s willingness to donate to a family member may be substantially higher than that to an unrelated recipient [5].

While previous studies have provided important insights, willingness to consider living kidney donation is likely shaped by multiple overlapping factors, including misconceptions about donor eligibility and outcomes, personal exposure to kidney disease or transplantation, financial considerations, and perceived barriers and motivators. Although many of these factors have been described individually, less is known about which beliefs, experiences, and concerns appear most closely tied to willingness when examined together within the same community-based sample. Identifying these patterns may help move living donation education beyond general awareness and toward the specific issues that influence whether community members would personally consider donation.

These questions may be particularly relevant in South Texas, a region with a large Hispanic population and a substantial burden of CKD and ESRD [9]. Prior studies have demonstrated more rapid CKD progression and worse kidney-related outcomes among Hispanic populations, as well as the influence of socioeconomic disadvantage on access to kidney transplantation and long-term outcomes [10–12]. San Antonio, Texas, represents one such community, with a high prevalence of kidney disease and significant socioeconomic diversity. Understanding attitudes, misconceptions, and concerns among community members regarding living kidney donation may help inform locally targeted educational and outreach initiatives aimed at expanding the living donor pool.

The purpose of this study was to evaluate factors associated with willingness to participate in living kidney donation in a diverse community-based population. Specifically, we sought to examine the relative contributions of living donation misconceptions, personal exposure to kidney disease and transplantation, financial considerations, and other perceived barriers and motivators to willingness to become a living kidney donor.

## Methods

### Study Design and Survey Development

A cross-sectional community-based survey was conducted to evaluate factors associated with willingness to participate in living kidney donation among adults in San Antonio, Texas. The survey was designed to assess attitudes toward living kidney donation, including willingness to become a living donor, personal exposure to kidney disease and transplantation, misconceptions regarding living donation, perceived barriers and motivators to donation, financial considerations, and demographic characteristics. The study was intended to examine the relative contributions of these factors to willingness to become a living kidney donor within a diverse community-based sample.

The questionnaire was developed specifically for this study after review of the relevant literature [4]. It has not previously been published as a complete survey instrument. The English-language version of the questionnaire is provided as a supplementary file. To ensure content validity, the survey questionnaire was reviewed by three transplant physicians with expertise in kidney transplantation and living donation prior to implementation. The English survey was translated into Spanish by a native Spanish speaker and then back into English by another. To ensure face validity, six volunteers, including two native Spanish speakers, completed the survey to assess the clarity of the questions.

The survey instrument used predominantly checkbox-based or binary responses to improve recruitment, reduce completion time, and facilitate administration on an Apple iPad using REDCap electronic data capture tools [13]. If preferred by participants, a QR code was also generated so that participants could scan and complete the survey on their own smartphones.

### Participant Recruitment and Data Collection

Survey data were collected from June 15 through August 30, 2024. Participants were directly approached and recruited using a community-based convenience sampling approach at multiple public locations across San Antonio, including parks on the weekends, community events, Independence Day celebrations, and bus stops. Parks were intentionally selected from different geographic regions and zip codes to improve the demographic diversity of the sample.

In-person enrollment was chosen because it allowed participation by individuals with limited internet access or who are less likely to respond to phone- or email-based surveys [14]. To reduce social desirability bias, participants completed the survey independently on an Apple iPad, and responses were entered directly into the REDCap cloud database without interviewer involvement during question completion [15].

To encourage participation and reduce nonresponse bias, non-monetary incentives such as bottled water and sodas were offered after survey completion [14].

### Ethical approval and consent to participate

This study was reviewed by the Institutional Review Board of The University of Texas Health Science Center at San Antonio (UT Health San Antonio) and was determined to be exempt under Exemption Category 2, 45 CFR 46.104(d)(2) [IRB protocol number: 00000473]. The study was determined to be exempt because the information obtained was recorded in a manner such that participants could not be identified, either directly or through identifiers linked to them. All participants received information about the study and participated voluntarily. Voluntary completion of the anonymous questionnaire constituted implied consent to participate. Under the exempt determination, signed or documented informed consent was not required. All methods and study procedures involving human participants were performed in accordance with the Declaration of Helsinki and applicable institutional and federal guidelines and regulations.

### Primary outcomes and survey variables

The primary outcome was willingness to participate in living kidney donation, assessed by the question “Would you consider becoming a living kidney donor?” This measure was intended to capture a general, unconditional willingness to become a living donor, rather than a willingness under specific circumstances or for a particular recipient.

Factors examined for association with willingness were grouped into four conceptual domains. Demographic variables included age, sex, race/ethnicity, education level, and household income. Misconception variables assessed beliefs regarding donor eligibility and outcomes, including whether only family members can donate, whether health insurance is required for donation, whether donors receive financial benefits, and whether donation increases the risk of kidney disease or shortens lifespan. Exposure variables assessed personal familiarity with kidney disease, transplantation, and living donation. Barrier and motivator variables included concerns regarding surgery, long-term health consequences, lost wages, religious considerations, reassurance regarding post-donation health, and altruistic motivations for donation.

Participants were permitted to select multiple responses for questions involving motivating and discouraging factors related to living donation.

### Statistical Analysis

Descriptive statistics were calculated for all survey variables. A total of 346 participants completed the survey; however, some participants did not complete all survey items. Percentages were therefore calculated using the number of respondents to each individual question. Analyses were conducted using complete cases for the variables included in each model. Participants who did not answer the primary outcome question were excluded from analyses of willingness. Given the very low level of missingness (0–2.9% across survey items), multiple imputation was not performed. The resulting sample size for each multivariable model is reported in the corresponding figure legend. Item-level missingness is summarized in Supplementary Table S1.

Descriptive statistics were used to summarize participant characteristics and survey responses. Categorical variables were summarized using frequencies and percentages.

Multivariable logistic regression was used to identify factors associated with willingness to participate in living kidney donation. To evaluate the relative contributions of different domains, a series of prespecified models were constructed. The variables were organized into domain-specific models to reflect conceptually distinct categories assessed by the survey: demographic and socioeconomic characteristics, personal exposure to kidney disease and transplantation, misconceptions regarding living donation, and perceived barriers and motivators. Within each domain-specific multivariable model, all variables in that domain were entered simultaneously, and each reported odds ratio was therefore adjusted for the other variables included in that model.

Model 1 evaluated associations between demographic factors and willingness to donate, including age, sex, race/ethnicity, education level, and household income.

Model 2 evaluated the association between personal exposure to kidney disease, transplantation, and living donation and willingness to donate.

Model 3 assessed the association between misconceptions about living donation and willingness to donate.

Model 4 evaluated the association between perceived barriers and motivators and willingness to donate.

Model 5 was an integrated model designed to evaluate the relative contributions of factors identified in the preceding analyses. Variables demonstrating significant associations based on domain-specific models were included in the final model to identify factors independently associated with willingness to participate in living kidney donation. Given the limited number of participants expressing willingness to donate, the integrated model was kept parsimonious to limit model overfitting.

Results are reported as odds ratios (ORs) with corresponding 95% confidence intervals (CIs). Statistical analyses were performed using STATA (College Station, TX).

## Results

### Participant Characteristics

A total of 346 participants completed the community survey. Demographic characteristics of the study population are summarized in Table 1. Because some participants did not complete all survey items, percentages for individual questions were calculated using the number of respondents to each specific item.

**Table 1.**
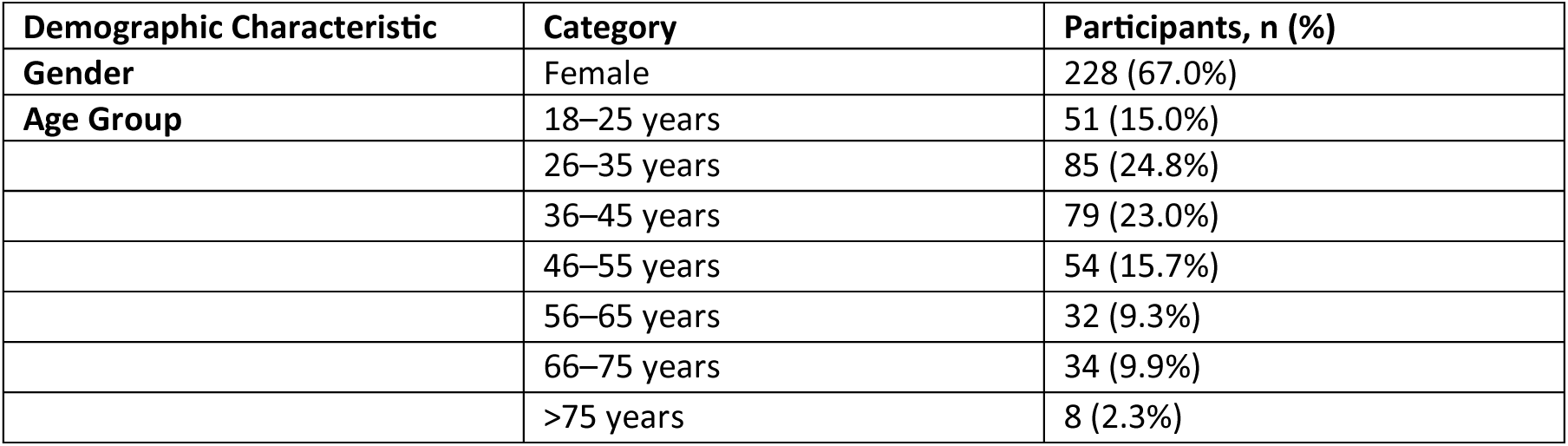
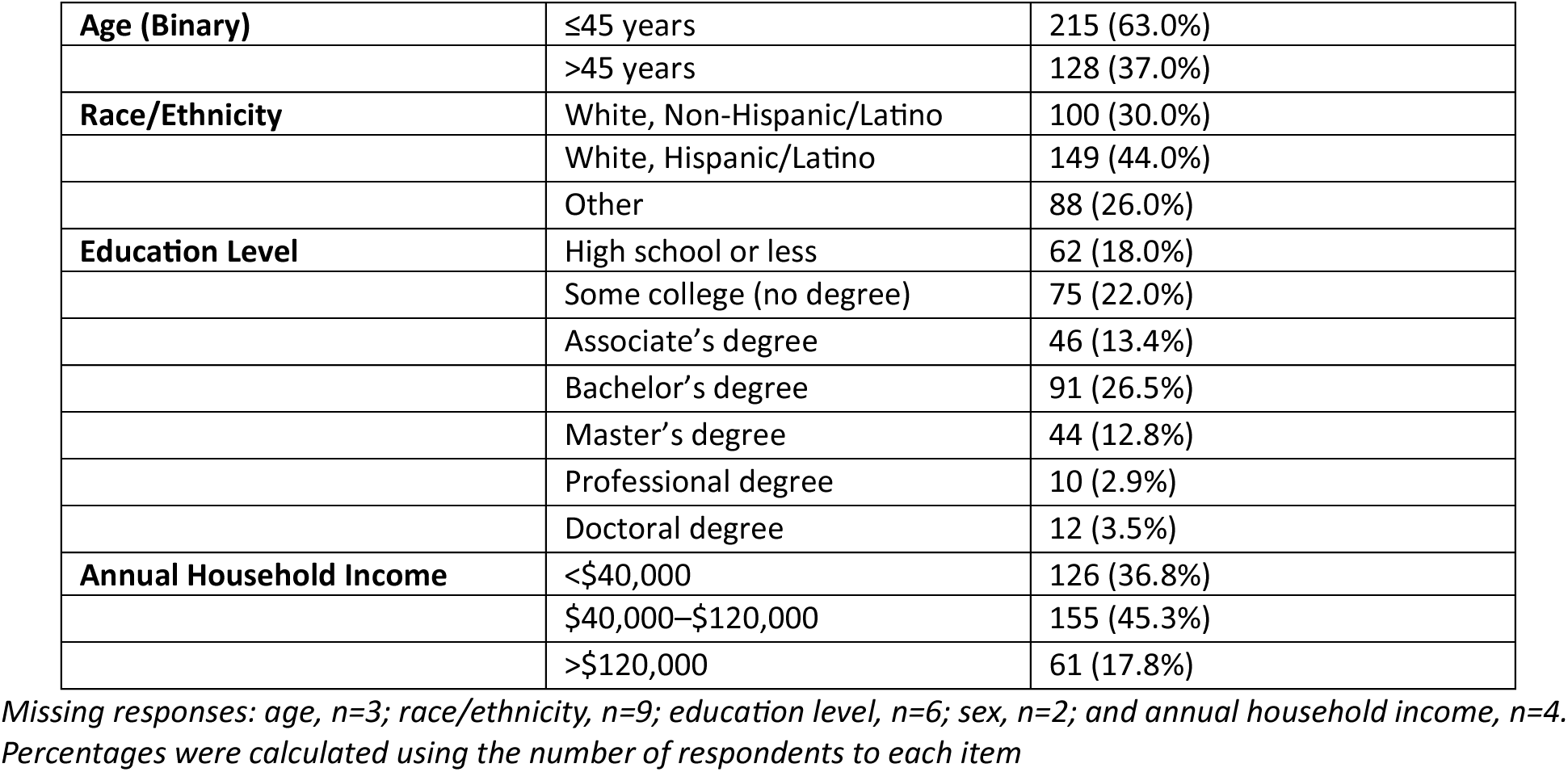
Demographic Characteristics of Survey Participants.

| Demographic Characteristic | Category | Participants, n (%) |
|---|---|---|
| **Gender** | Female | 228 (67.0%) |
| **Age Group** | 18–25 years | 51 (15.0%) |
|  | 26–35 years | 85 (24.8%) |
|  | 36–45 years | 79 (23.0%) |
|  | 46–55 years | 54 (15.7%) |
|  | 56–65 years | 32 (9.3%) |
|  | 66–75 years | 34 (9.9%) |
|  | >75 years | 8 (2.3%) |
| **Age (Binary)** | ≤45 years | 215 (63.0%) |
|  | >45 years | 128 (37.0%) |
| **Race/Ethnicity** | White, Non-Hispanic/Latino | 100 (30.0%) |
|  | White, Hispanic/Latino | 149 (44.0%) |
|  | Other | 88 (26.0%) |
| **Education Level** | High school or less | 62 (18.0%) |
|  | Some college (no degree) | 75 (22.0%) |
|  | Associate’s degree | 46 (13.4%) |
|  | Bachelor’s degree | 91 (26.5%) |
|  | Master’s degree | 44 (12.8%) |
|  | Professional degree | 10 (2.9%) |
|  | Doctoral degree | 12 (3.5%) |
| **Annual Household Income** | <\$40,000 | 126 (36.8%) |
|  | \$40,000–\$120,000 | 155 (45.3%) |
|  | >\$120,000 | 61 (17.8%) |

*Missing responses: age, n=3; race/ethnicity, n=9; education level, n=6; sex, n=2; and annual household income, n=4. Percentages were calculated using the number of respondents to each item*

Of the 343 participants who answered the primary outcome question, 68 (19.8%) expressed willingness to become a living kidney donor

### Awareness about living donation, general and conditional willingness to participate in living donation

Although 83.8% of participants were aware that a healthy individual can donate one kidney as a living donor, understanding of the role of kidney transplantation in treating kidney failure was more limited. Among 340 participants who answered the question regarding the best treatment for kidney failure, only 136 (40.0%) identified kidney transplantation as the best treatment, while 116 (34.1%) responded that they did not know, 49 (14.4%) selected dialysis, and 39 (11.5%) believed that dialysis and transplantation were equivalent.

When asked to whom they would be willing to donate a kidney, participants most selected family members or close friends (51.7%). Fewer participants reported a willingness to donate to anyone in need (28.6%) or to a person with a compelling personal story (21.4%). Smaller proportions reported willingness to donate to a sick child (17.1%) or to a young child’s parent (11.3%). Overall, 64 participants (18.5%) indicated that they would not donate a kidney. Because participants could select more than one recipient category, responses were not mutually exclusive.

### Demographic and socioeconomic factors associated with willingness to donate

Demographic and socioeconomic factors associated with willingness to become a living kidney donor are summarized in Figure 1. After adjustment for age, sex, ethnicity, education, employment status, and household income, no independent demographic or socioeconomic variables demonstrated statistically significant associations with willingness to become a living donor (Figure 1)

**Figure 1.**
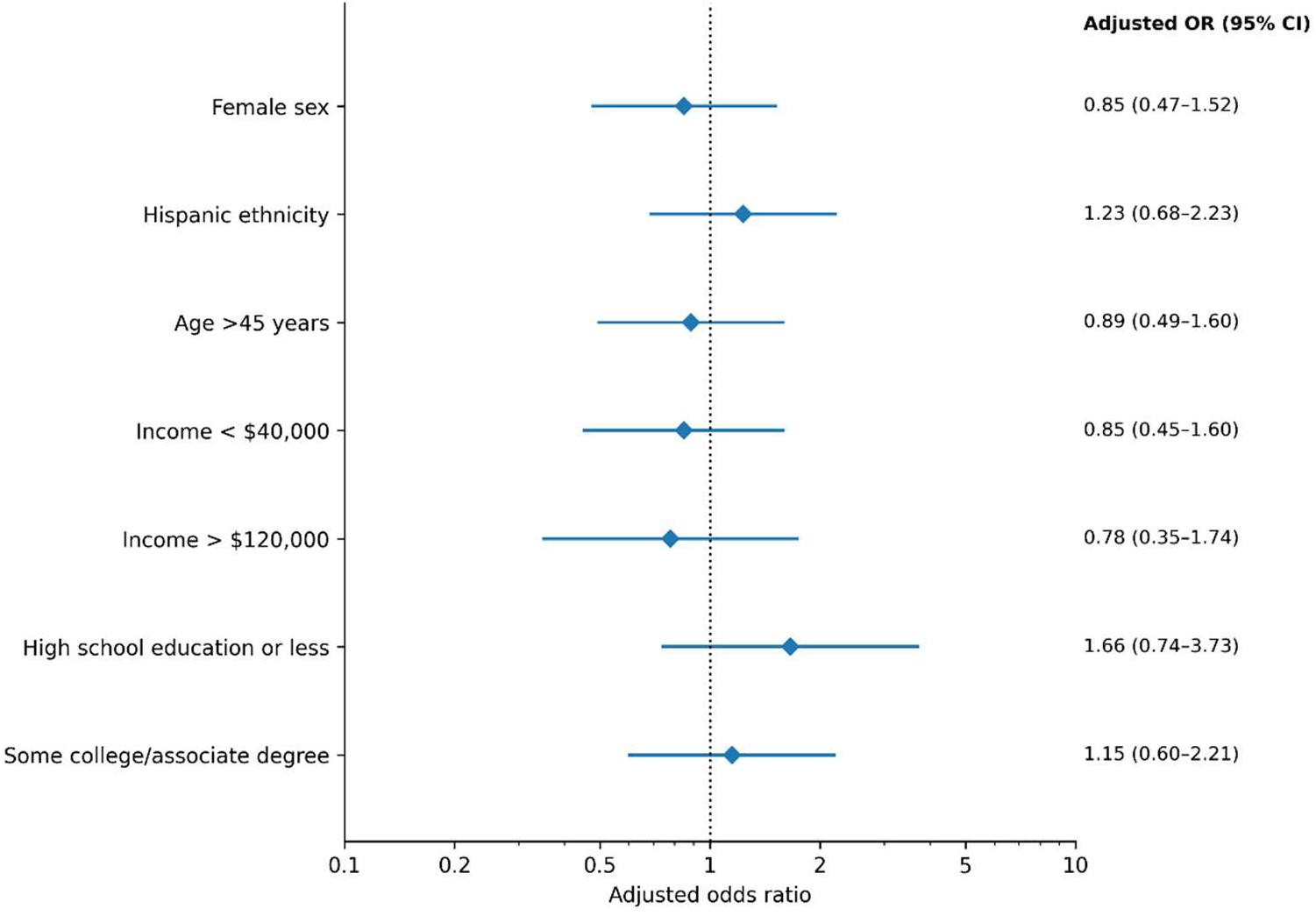
Demographic and socioeconomic factors associated with willingness to become a living kidney donor (Model 1, n=328) Reference groups were male sex, non-Hispanic ethnicity, age ≤45 years, annual household income $40,000–$120,000, and bachelor’s degree or higher

### Exposure to transplantation

Many participants reported prior exposure to kidney disease and transplantation through family members or acquaintances. Approximately 17.1% of participants reported knowing someone currently receiving dialysis, 22.8% knew someone who had undergone kidney transplantation, and 21.8% knew someone who had donated a kidney. Familiarity with an individual currently receiving dialysis was not associated with willingness to become a living donor. In contrast, familiarity with a transplant recipient was associated with lower willingness to participate in living donation, whereas familiarity with a living kidney donor was associated with greater willingness to become a living donor (Figure 2)

**Figure 2.**
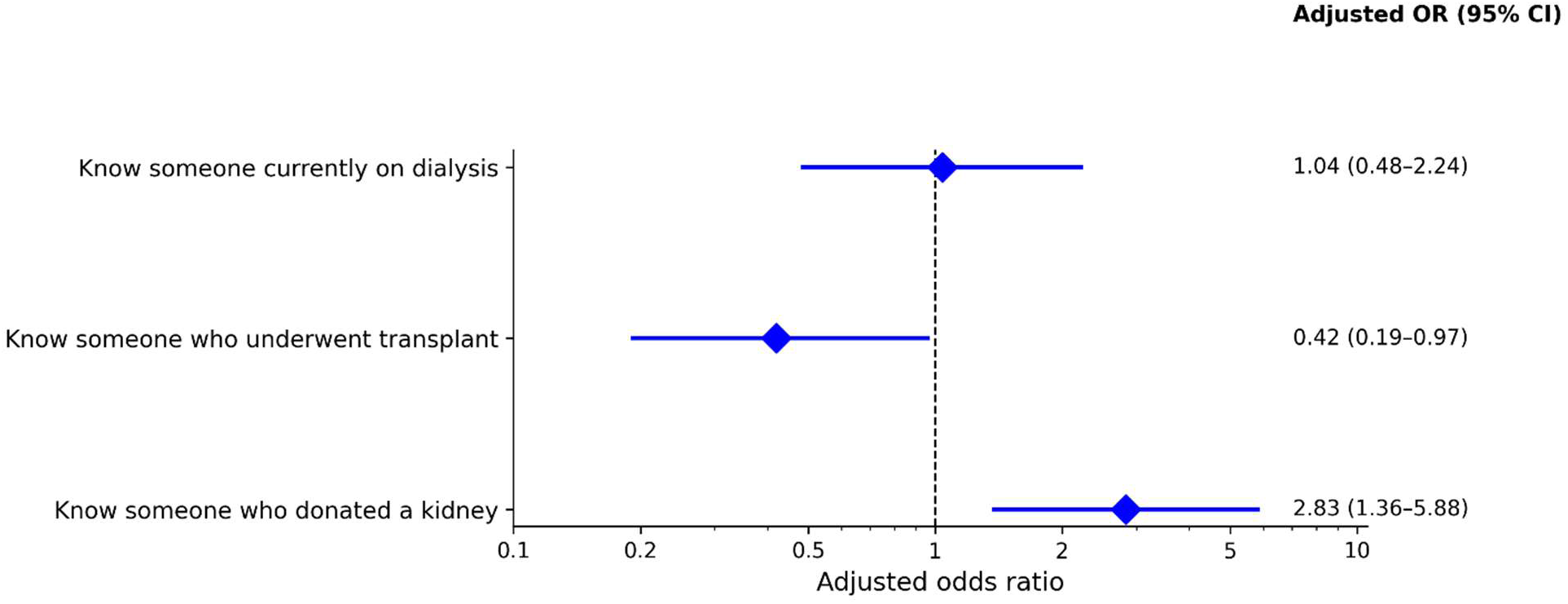
Association between personal exposure to kidney disease, transplantation and living donation and willingness to donate (Model 2, N=337) For each exposure, the reference group was participants who did not report the corresponding exposure.

### Knowledge and Misconceptions Regarding Living Donation

Several misconceptions regarding living donation and transplantation were identified. The two most common misconceptions were the beliefs that donating a kidney increases the donor’s future risk of kidney disease (22.4%) and that donors are responsible for all medical costs associated with donation (22.2%). Additional misconceptions included beliefs that organ donors receive financial compensation for donation (12.6%), that living donation shortens lifespan (12.8%), and that women who donate a kidney cannot later become pregnant (8%). Awareness regarding living kidney donation was generally high. Most participants (84.6%) correctly stated that donors must generally be healthy before donation. Most participants also correctly recognized that organ donation is not limited to family members alone (90.0%) and that medical insurance is not required to become an organ donor (87.5%).

In an exploratory analysis, the cumulative number of living donation misconceptions was not associated with willingness to donate (OR 0.99 per additional misconception, 95% CI 0.79–1.24)

None of the individual misconception variables were independently associated with willingness to become a living donor (Figure 3).

**Figure 3.**
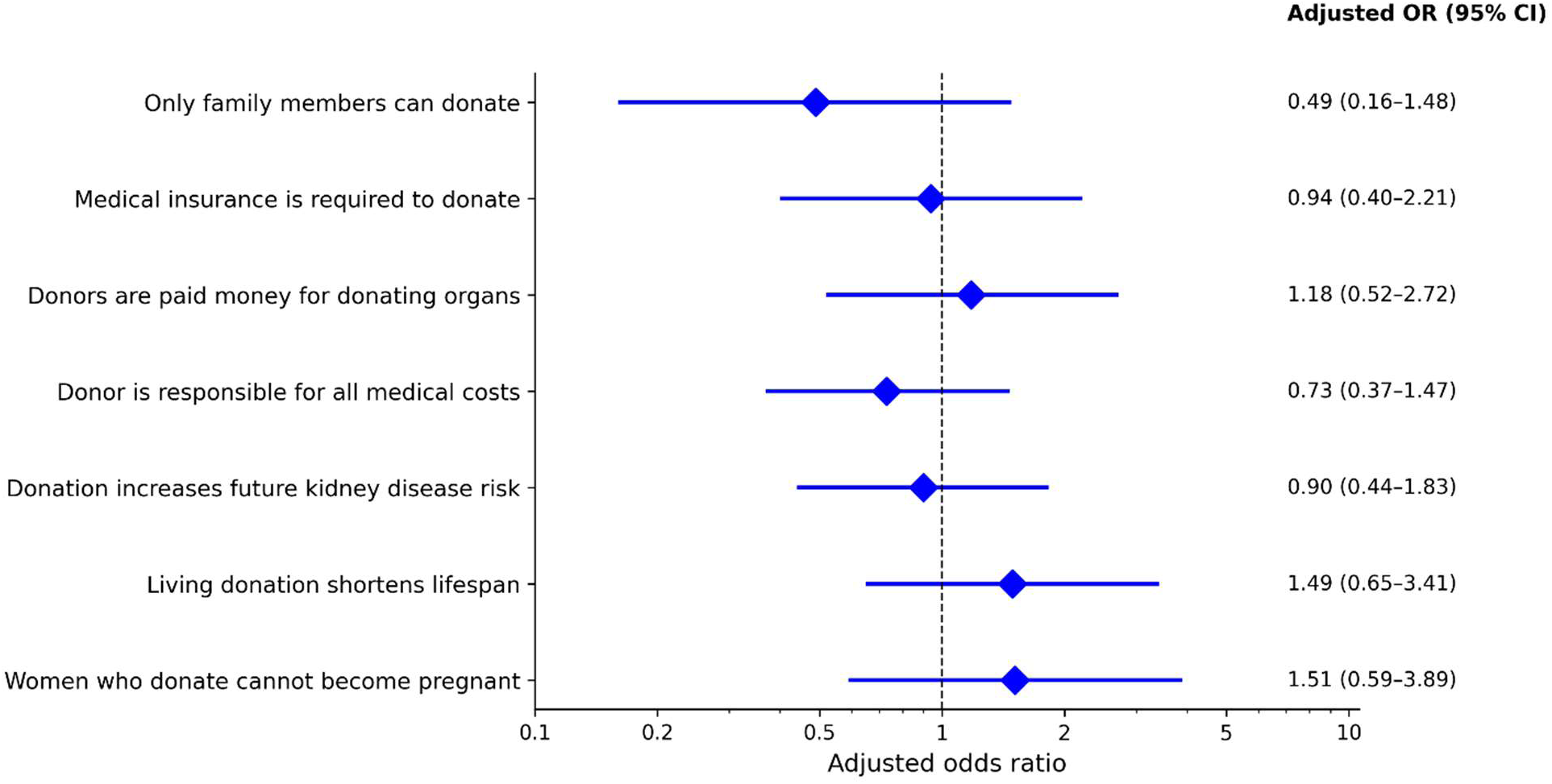
Association of living donation misconceptions with willingness to donate (Model 3, N=336) For each misconception, the reference group was participants who did not endorse the listed misconception

### Motivating and discouraging factors associated with willingness to donate

Participants identified several discouraging and motivating factors related to living kidney donation. Concerns about long-term health complications were the most reported discouraging factor (40.5%), followed by fear of surgery (28.3%). Additional concerns included losing wages during recovery (18.8%) and religious or personal beliefs (8.4%).

Among motivating factors, helping someone in need was the most frequently selected response (46.8%), followed by reassurance regarding the ability to live a healthy life after donation (34.1%). Financial compensation (17.9%) and assurance of health insurance coverage (15.6%) were less frequently selected motivating factors.

A separate multivariable logistic regression model evaluating motivating and discouraging factors associated with willingness to become a living donor is summarized in Figure 4. Participants who identified “helping someone in need” as a motivating factor demonstrated significantly higher odds of expressing willingness to become a living donor (aOR 2.97, 95% CI 1.64–5.40, p<0.001). Reassurance regarding the ability to live a healthy life following donation was also independently associated with greater willingness to donate (aOR 2.44, 95% CI 1.20–4.97).

**Figure 4:**
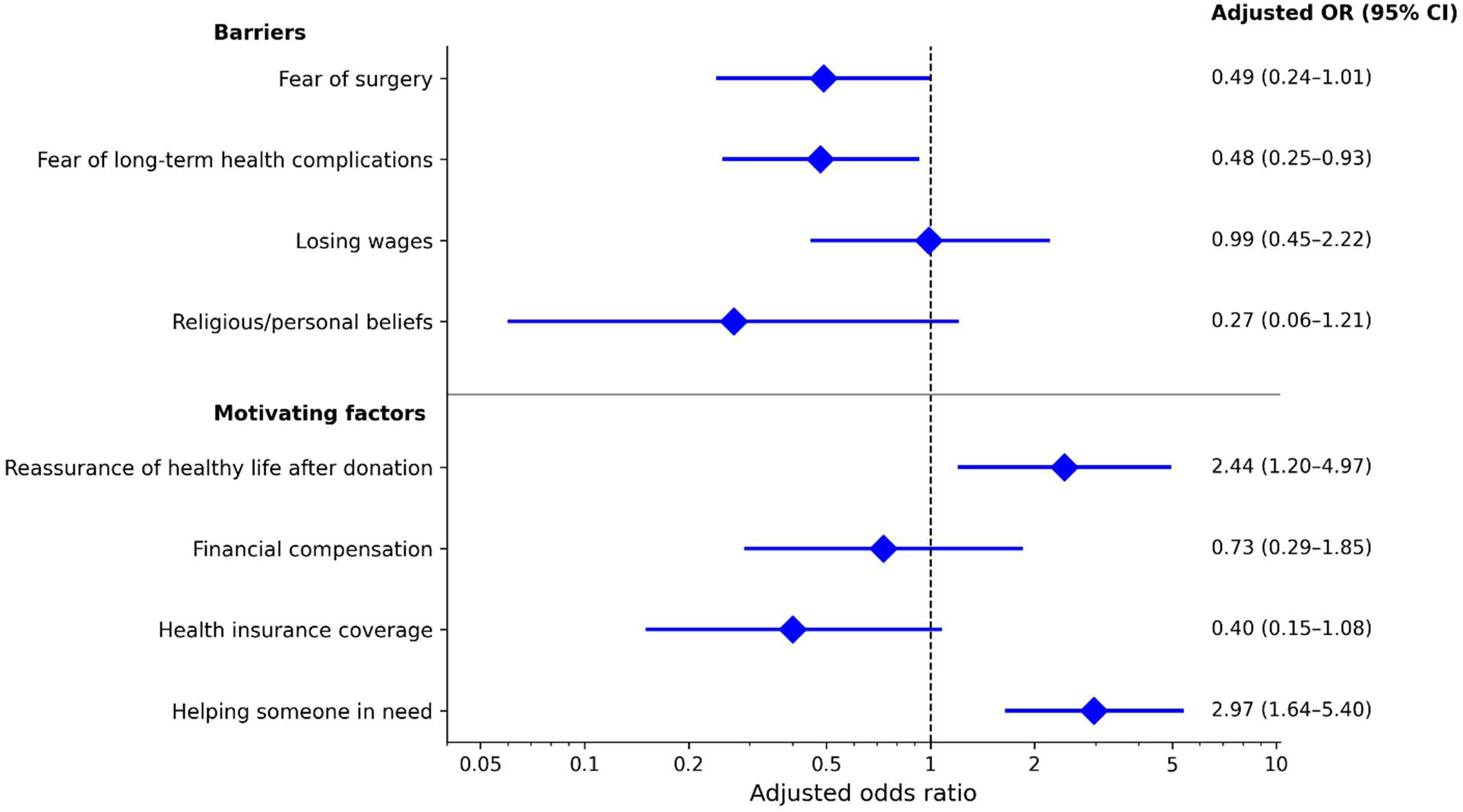
Motivating and discouraging factors associated with willingness to donate (Model 4, N=339) For each barrier or motivator, the reference group was participants who did not select the listed factor.

Conversely, concern regarding long-term health complications was independently associated with lower willingness to become a living donor (aOR 0.48, 95% CI 0.25–0.93, p=0.03). Fear of surgery demonstrated a borderline negative association with donation willingness (aOR 0.49, 95% CI 0.24–1.01, p=0.053) (Figure 4)

Exploratory subgroup analyses showed that perceived barriers and motivators differed by age for fear of surgery (p=0.033), concern about lost wages (p=0.003), and financial compensation as a motivator (p=0.002). Religious or personal beliefs as a discouraging factor differed by ethnicity (p=0.002), and fear of surgery differed by sex (p=0.044). No significant differences in perceived barriers or motivators were observed by race.

### Financial compensation and willingness to donate

Financial compensation was further evaluated by asking participants what amount, if any, would motivate them to consider living kidney donation. Among 336 respondents who answered this question, 171 (50.9%) reported that no amount of money would motivate them to donate, while 165 (49.1%) selected a monetary compensation amount. The most selected threshold was $100,000 (55/336, 16.4%), followed by $50,000 (45/336, 13.4%), $20,000 (34/336, 10.1%), and $10,000 (31/336, 9.2%). Among 275 participants who were not initially willing to become living donors, 125 (45.5%) selected a compensation amount, 139 (50.5%) reported that no amount would motivate them to donate, and 11 (4.0%) skipped the question or provided no response. In a descriptive cross-tabulation, selection of any monetary compensation amount differed significantly by age (p<0.001) but not by household income (p=0.91).

Among respondents who answered the compensation-amount question, the proportion selecting any monetary amount declined progressively with age, from 80.0% among participants aged 18–25 years to 14.3% among those older than 75 years (p<0.001). In contrast, selection of a monetary compensation amount did not differ meaningfully across household income categories (p=0.8).

### Relative contribution of significant factors associated with willingness to donate

Variables demonstrating significant associations in the domain-specific analyses were incorporated into an integrated multivariable model to assess their relative contributions to willingness to participate in living kidney donation. These included familiarity with a transplant recipient, familiarity with a living kidney donor, fear of long-term health complications, helping someone in need, and reassurance of a healthy life after donation. In the integrated model, familiarity with a living kidney donor (OR 2.82, 95% CI 1.33–5.95) and helping someone in need (OR 2.48, 95% CI 1.46–4.75, p=0.001) were independently associated with greater willingness to become a living donor. In contrast, familiarity with a transplant recipient (OR 0.42, 95% CI 0.19–0.96, p=0.03) and concern regarding long-term health complications (OR 0.48, 95% CI 0.26–0.90, p=0.027) were associated with lower willingness. Reassurance of a healthy life after donation demonstrated a borderline positive association with willingness to donate (OR 1.64, 95% CI 0.89–3.02, p=0.080) (Figure 5)

**Figure 5.**
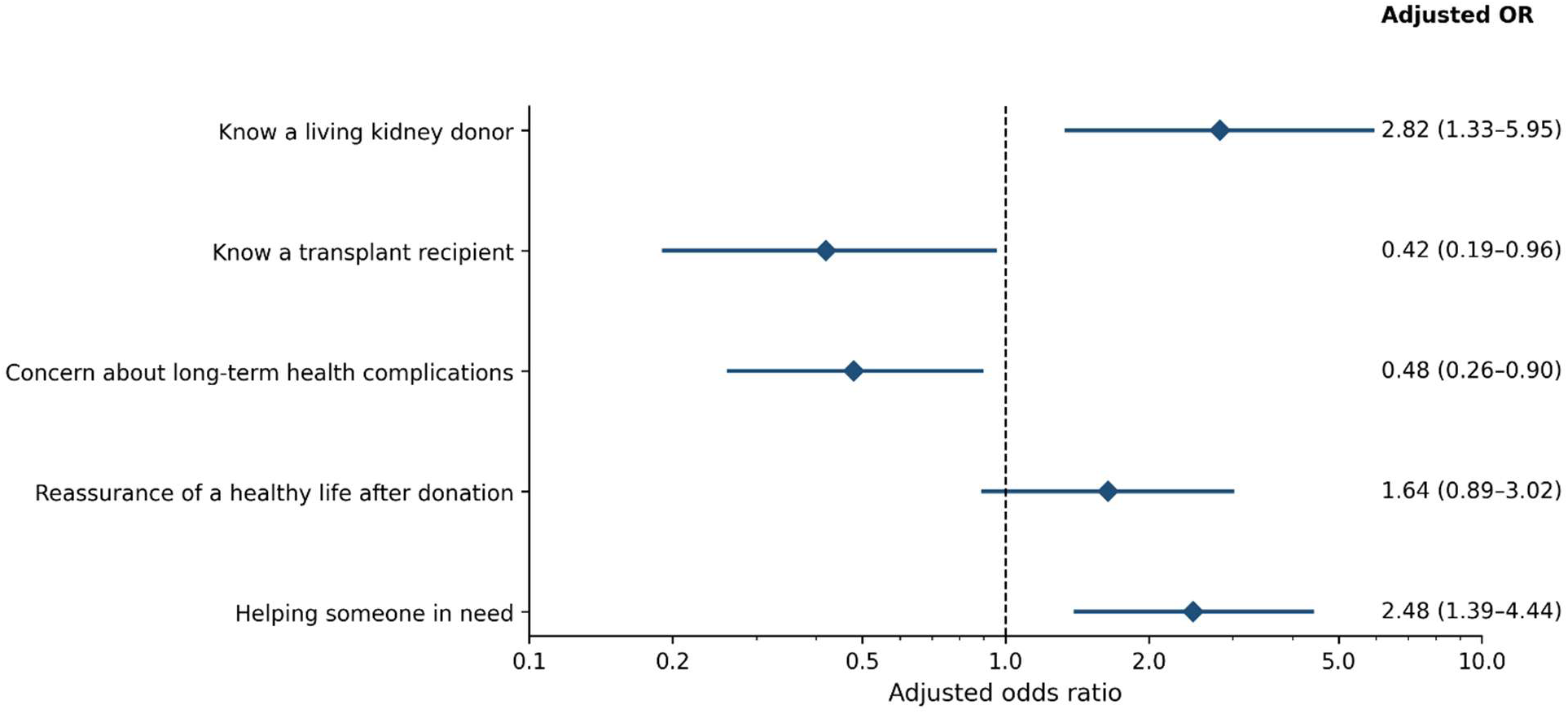
Relative Contributions of Exposure, Barriers, and Motivators Associated with Willingness to Become a Living Kidney Donor (Model 5, N=339) For each factor, the reference group was participants who did not report or select the corresponding factor.

## Discussion

Only one in five participants expressed willingness to become a living kidney donor. Willingness was associated with personal exposure, perceived donor safety, and motivation to help others. Familiarity with a living kidney donor and identifying helping someone in need as a motivator were associated with greater willingness, whereas familiarity with a transplant recipient and concern regarding long-term health complications were associated with lower willingness. Although most participants knew that a healthy person could donate one kidney, only 40% knew that kidney transplantation was the best treatment for ESRD, suggesting that awareness of living donation alone may be insufficient to translate into willingness to donate. Demographic and socioeconomic factors, as well as individual misconceptions about living donation, were not independently associated with willingness. Financial compensation was not independently associated with willingness in the motivator model, although nearly half of initially unwilling respondents selected a compensation amount that could potentially motivate donation, with younger participants more likely to do so.

### Gap between awareness and willingness to participate in living kidney donation

A major finding of this study was the gap between awareness of living donation and willingness to personally participate. In our cohort, 83.8% of participants knew that a healthy person could donate one kidney while living, which was only modestly lower than the 91% awareness reported in a recent national survey [5]. However, willingness to donate was substantially lower in our study. Only 19.6% of participants expressed willingness to become a living donor, compared with prior national surveys in which approximately half of respondents reported willingness to donate to a stranger [4,5]. Even willingness to donate to a family member or close friend was lower in our cohort at 51.7%, compared with approximately 82% in the national survey [5]. These differences are difficult to explain by awareness of living donation alone.

One possible explanation is that awareness of living donation may not be accompanied by an understanding of the benefit of kidney transplantation itself. Only 40.0% of respondents in our study identified kidney transplantation as the best treatment for eligible patients with kidney failure, while more than one-third reported that they did not know, and another quarter selected dialysis or believed dialysis and transplantation were equivalent. To our knowledge, perception of transplantation as the preferred treatment for eligible patients with kidney failure has not been routinely assessed in prior public surveys and may represent a novel and actionable finding from this study. This gap is clinically important. Prior studies have shown that five-year survival after dialysis initiation is only 40%, yet dialysis patients often overestimate their long-term prognosis [16]. At the same time, discussions of dialysis are often dominated by quality-of-life concerns such as symptom burden, fatigue, cramps, dietary restrictions, and the time commitment of hemodialysis [17]. Without a clear understanding that kidney transplantation improves both survival and quality of life compared with remaining on dialysis [18], and that living donor transplantation offers additional advantages over deceased donor transplantation [3], members of the public may not view living donation as urgent, necessary, or personally meaningful. Community education may therefore need to explain not only that living donation is possible, but also why transplantation is the preferred treatment for eligible patients with kidney failure.

The gap between awareness and willingness may also reflect regional demographics, survey methodology, sampling approaches, or other unmeasured factors.

### Personal Exposure to a Living Kidney Donor and Transplant Recipient

There is some evidence to suggest that personal exposure to kidney transplantation may influence public attitudes toward living donation. For example, in the Latin American population living in Spain, exposure to kidney transplantation was associated with greater willingness to become a living donor [19]. However, that study did not distinguish whether exposure occurred through knowing a living kidney donor, knowing a transplant recipient, or both. Our findings suggest that this distinction may be important. Familiarity with a living kidney donor was independently associated with greater willingness to participate in living donation, whereas familiarity with a transplant recipient was associated with lower willingness. These findings suggest that “exposure to transplantation” may not be a uniform experience.

Knowing a living donor may normalize donation and provide reassurance that donors can recover and live healthy lives after nephrectomy. One possible explanation for the association between familiarity with a transplant recipient and lower willingness to donate is perceived personal risk. In South Texas, where diabetes and kidney disease are common and may cluster within families [9], knowing a transplant recipient may make participants more aware of their own potential risk for developing CKD. Because diabetes is a leading cause of kidney failure [20] and widely prevalent in San Antonio [21], individuals who know a transplant recipient personally may be more likely to view kidney disease as something that could affect themselves or their family members. This perceived vulnerability could reduce willingness to donate a kidney, even if the individual supports living donation in principle. Since our survey did not directly assess diabetes history, family history of kidney disease, or perceived personal risk, this explanation remains speculative and should be evaluated in future studies.

### Altruism, donor health concerns and reassurance after donation

Altruistic motivation and donor health concerns appeared to represent two of the most important and opposing influences on willingness to participate in living kidney donation. In our study, identifying helping someone in need as a motivator was associated with willingness to donate, whereas fear of long-term health complications was independently associated with lower willingness to donate. Reassurance regarding the ability to live a healthy life after donation was also positively associated with willingness, although this association was attenuated in the final integrated model. Together, these findings suggest that willingness to donate may depend on both the desire to help another person and confidence that donation will not result in unacceptable long-term personal harm. This pattern is consistent with prior literature showing that altruism is a central motivator for living donation [22], whereas concerns regarding donor safety and long-term health outcomes remain among the most important barriers [4]. Interestingly, individual misconceptions were not independently associated with willingness in multivariable analysis, suggesting that donation attitudes may be shaped less by isolated factual errors and more by broader perceptions of long-term health concerns and well-being. These findings support the need for educational interventions that not only appeal to altruistic motivations but also provide concrete reassurance about donor evaluation, recovery, long-term kidney health, pregnancy after donation, and ongoing post-donation follow-up.

### Financial Compensation and Willingness to Donate

Financial compensation represented a more complex finding. In the multivariable model of motivators and barriers, financial compensation was not independently associated with willingness to become a living donor. However, responses to the hypothetical financial incentives question suggest that financial considerations may still influence donation attitudes for a substantial subset of participants. Nearly half of all respondents who answered the question selected some compensation amount that could motivate donation, and among participants who were not initially willing to become living donors, 45.5% selected a monetary threshold. At the same time, 50.5% of initially unwilling participants reported that no amount of money would motivate them to donate, indicating that financial compensation alone would not address unwillingness for many individuals. Notably, the selection of a compensation amount varied strongly by age but not by household income, declining from 80.0% among participants aged 18–25 years to 14.3% among those older than 75 years, with no meaningful gradient across income categories. This suggests that openness to compensation may reflect age-related factors such as financial insecurity, student debt, lost wages, risk tolerance, or life stage rather than income alone, but the survey was not designed to distinguish among those reasons. Although direct payment for donated organs remains prohibited under the National Organ Transplant Act [23], the persistent stagnation of living donation rates has prompted renewed policy discussion of non-cash incentives, such as tax credits or deductions for donation-related costs, as a legally and ethically permissible middle ground between prohibition and payment [24]. Until there is a change in policy, it remains relevant to address financial concerns among potential donors directly and transparently through legally permissible donor protections, including reimbursement for travel, lost wages, dependent care, and other donation-related expenses, which are often available through the National Living Donor Assistance Center [25].

### Implications for Living Donation Outreach

These findings have several practical implications for community-based living donation outreach. Educational efforts should move beyond basic awareness that living donation is possible and should also explain why kidney transplantation is generally the preferred treatment for eligible patients with kidney failure. ESRD treated with dialysis should not be framed only as a quality-of-life burden involving fatigue, dietary restriction, time commitment, and treatment inconvenience; the public should also understand the high mortality associated with long-term dialysis and the survival and quality-of-life benefits associated with kidney transplantation, particularly living donor transplantation. Outreach efforts should also include living donor stories and use donor champions, because familiarity with a living donor was associated with willingness to donate and may help normalize donation, reduce fear, and provide reassurance about recovery and long-term health after nephrectomy. At the same time, educational programs should directly address concerns about long-term donor safety, kidney function after donation, pregnancy after donation, and post-donation follow-up. Financial issues should also be discussed transparently, including existing protections, reimbursement programs, lost-wage concerns, and insurance-related concerns, especially because financial compensation appeared relevant to a substantial subset of younger respondents. Finally, because this study was conducted in San Antonio, outreach should be locally tailored and culturally responsive, with messaging that reflects the burden of diabetes, kidney disease, and transplantation in this community.

### Limitations

This study has several limitations. First, participants were recruited using a community-based convenience sampling approach in San Antonio, which may limit generalizability to other regions or populations. However, surveys were administered in person at multiple public locations and zip codes to improve demographic diversity, allowing participation from individuals who may be less likely to respond to online or telephone-based surveys. Second, the cross-sectional design limits causal inference; associations between exposure, concerns, motivators, and willingness should not be interpreted as evidence that these factors directly cause willingness or unwillingness to donate. Third, the primary outcome was self-reported hypothetical willingness to become a living donor rather than actual donor registration, evaluation, or donation behavior. Fourth, the survey utilized primarily binary response options rather than Likert-scale measures. While this approach facilitated rapid completion in community settings, reduced respondent burden, and was practical during in-person summer recruitment in San Antonio, it may have limited the ability to capture gradations in attitudes and beliefs regarding living donation. Finally, because only a minority of participants expressed willingness to become living donors, the number of outcome events was limited, potentially reducing the power to detect weaker associations.

## Conclusion

In conclusion, this community-based survey suggests that willingness to participate in living kidney donation is shaped by more than awareness alone. Although most participants knew that living donation is possible, fewer understood the role of kidney transplantation as the preferred treatment for eligible patients with kidney failure, and only one in five expressed general willingness to become a living donor. Willingness was associated with familiarity with a living kidney donor, altruistic motivation, donor safety concerns, and financial considerations, rather than with demographic factors or isolated misconceptions alone. These findings suggest that efforts to expand the living donor pool should move beyond basic awareness campaigns and instead combine education about the survival and quality-of-life benefits of transplantation, reassurance regarding long-term donor safety, real stories from living donors, and transparent discussion of financial barriers and protections. Such locally tailored approaches may be particularly important in communities with a high burden of diabetes and kidney disease, as well as socioeconomic diversity.

## Data Availability

All data produced in the present study are available upon reasonable request to the authors

## Declarations

### Ethics statement and consent to participate

The study was submitted to the UT Health San Antonio Institutional Review Board. The study (STUDY 00000477) was determined to be exempt by the IRB, and written informed consent was therefore not required as per federal regulations (45 CFR 46.104(d)(2).

### Consent for publication

Not applicable

### Availability of data and materials

The datasets used and/or analyzed during the current study are available from the corresponding author upon reasonable request.

### Authors’ contributions

SM conceived the study, designed the study, acquired the data, analyzed the data and drafted the manuscript. ET contributed to the survey instrument design and revised the manuscript. Both authors agree with the submitted version of the manuscript.

## Acknowledgments

None

---

**Supplementary table S1.**
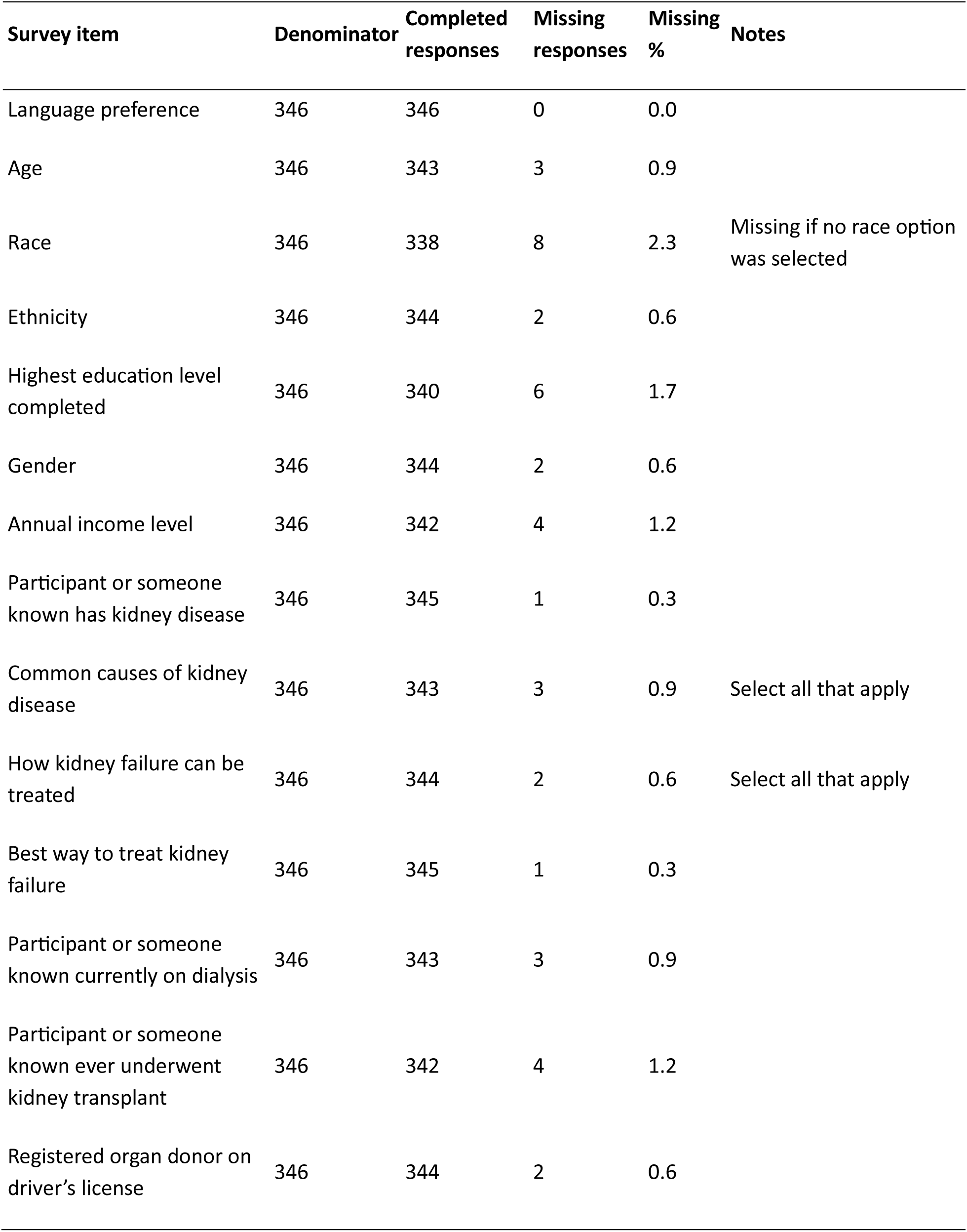
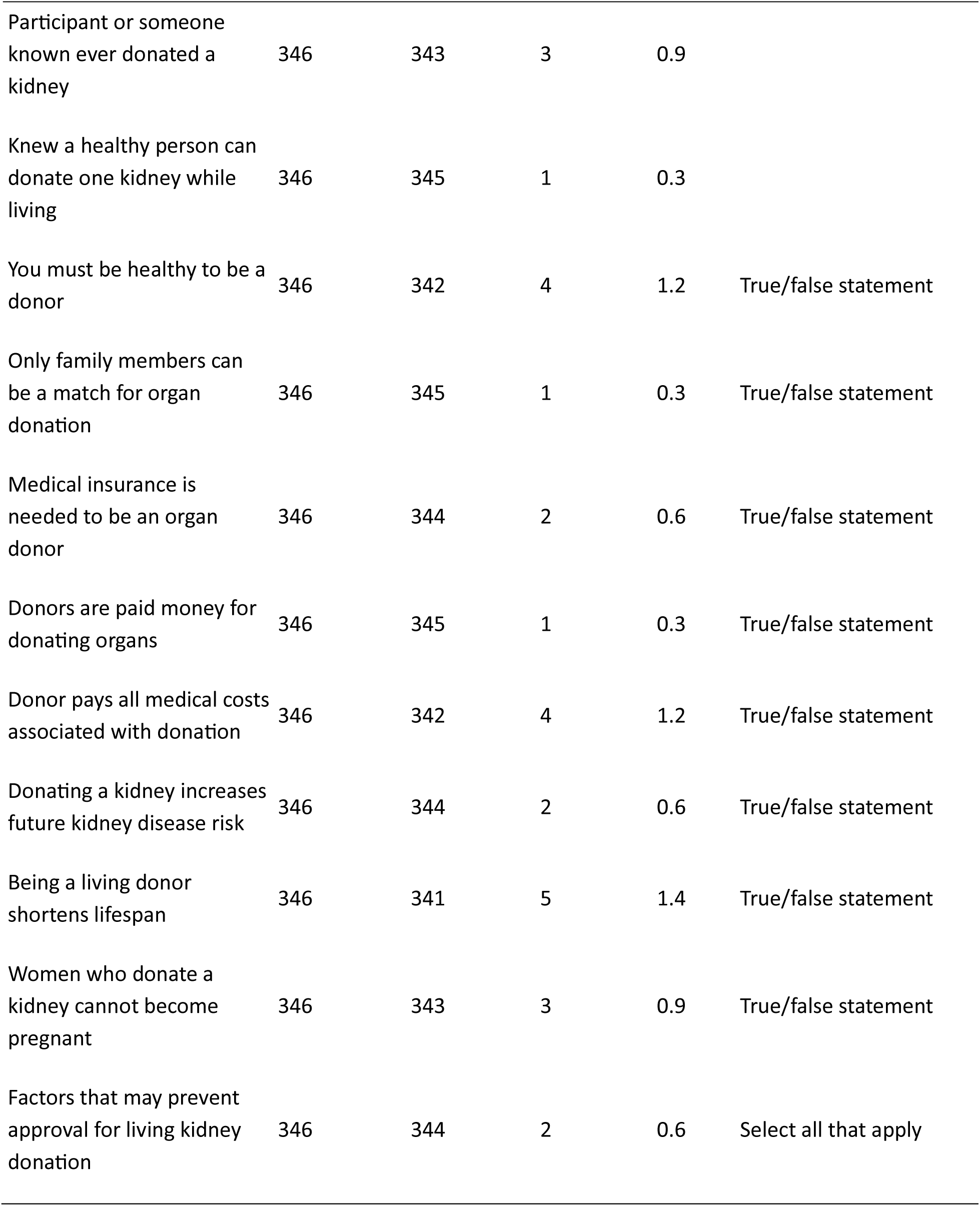
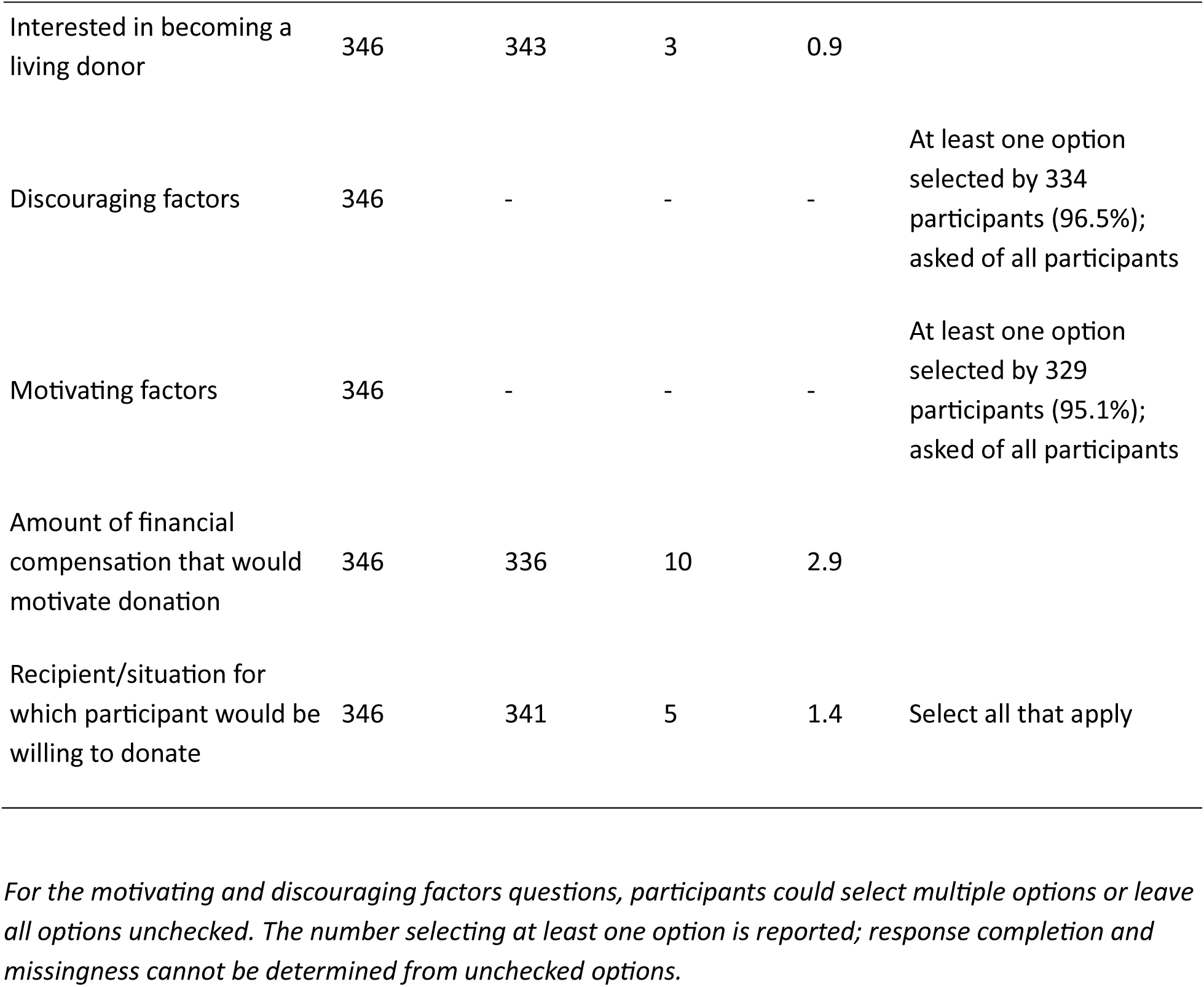
Missing responses across the questionnaire.

| **Survey item** | **Denominator** | **Completed responses** | **Missing responses** | **Missing %** | **Notes** |
|---|---|---|---|---|---|
| Language preference | 346 | 346 | 0 | 0.0 |  |
| Age | 346 | 343 | 3 | 0.9 |  |
| Race | 346 | 338 | 8 | 2.3 | Missing if no race option was selected |
| Ethnicity | 346 | 344 | 2 | 0.6 |  |
| Highest education level completed | 346 | 340 | 6 | 1.7 |  |
| Gender | 346 | 344 | 2 | 0.6 |  |
| Annual income level | 346 | 342 | 4 | 1.2 |  |
| Participant or someone known has kidney disease | 346 | 345 | 1 | 0.3 |  |
| Common causes of kidney disease | 346 | 343 | 3 | 0.9 | Select all that apply |
| How kidney failure can be treated | 346 | 344 | 2 | 0.6 | Select all that apply |
| Best way to treat kidney failure | 346 | 345 | 1 | 0.3 |  |
| Participant or someone known currently on dialysis | 346 | 343 | 3 | 0.9 |  |
| Participant or someone known ever underwent kidney transplant | 346 | 342 | 4 | 1.2 |  |
| Registered organ donor on driver’s license | 346 | 344 | 2 | 0.6 |  |
| Participant or someone known ever donated a kidney | 346 | 343 | 3 | 0.9 |  |
| Knew a healthy person can donate one kidney while living | 346 | 345 | 1 | 0.3 |  |
| You must be healthy to be a donor | 346 | 342 | 4 | 1.2 | True/false statement |
| Only family members can be a match for organ donation | 346 | 345 | 1 | 0.3 | True/false statement |
| Medical insurance is needed to be an organ donor | 346 | 344 | 2 | 0.6 | True/false statement |
| Donors are paid money for donating organs | 346 | 345 | 1 | 0.3 | True/false statement |
| Donor pays all medical costs associated with donation | 346 | 342 | 4 | 1.2 | True/false statement |
| Donating a kidney increases future kidney disease risk | 346 | 344 | 2 | 0.6 | True/false statement |
| Being a living donor shortens lifespan | 346 | 341 | 5 | 1.4 | True/false statement |
| Women who donate a kidney cannot become pregnant | 346 | 343 | 3 | 0.9 | True/false statement |
| Factors that may prevent approval for living kidney donation | 346 | 344 | 2 | 0.6 | Select all that apply |
| Interested in becoming a living donor | 346 | 343 | 3 | 0.9 |  |
| Discouraging factors | 346 | - | - | - | At least one option selected by 334 participants (96.5%); asked of all participants |
| Motivating factors | 346 | - | - | - | At least one option selected by 329 participants (95.1%); asked of all participants |
| Amount of financial compensation that would motivate donation | 346 | 336 | 10 | 2.9 |  |
| Recipient/situation for which participant would be willing to donate | 346 | 341 | 5 | 1.4 | Select all that apply |

*For the motivating and discouraging factors questions, participants could select multiple options or leave all options unchecked. The number selecting at least one option is reported; response completion and missingness cannot be determined from unchecked options.*

